# Prediction of Spreads of COVID-19 in India from Current Trend

**DOI:** 10.1101/2020.05.01.20087460

**Authors:** Himanshu Shekhar

**Affiliations:** O/o DG(ACE), (DRDO), Pashan, Pune–411021

**Keywords:** COVID-19, India, Modeling, Current Trend, Prediction, Infection, Maharastra, Gujarat, Delhi

## Abstract

The article describe modelling efforts for evaluating the current level of COVID-19 infections in India, using exponential model. The Data from 15 march 2020 to 30 April 2020 are used for validating the model, where intrinsic rise rate is kept constant. It is observed that some states of India, like MAharastra, Gujarat and Delhi have a much higher daily infection cases. This is modelled by assuming an initial higher infections, keeping rise rate same. The sudden outbursts are captured using offset of values for these three states. Data from other states like Madhya Pradesh, Uttar Pradesh and Rajasthan are also analysed and they are found to be following the same constants as India is following. Worldwide, many attempts are made to predict outburst of COVID-19 and in the model, described in this paper, turning point is not predicted, as cases in India are still rising. The developed model is based on daily confirmed infections and not on cumulative infections and rationalization is carried out for the population of various regions, while predicting infections for various states. Assigning a decay constant at this stage will be a premature exercise and keeping that in mind, exponential model predicts that India will attain 1 lakh case by 15 May 2020. The figure of 2 lakh and 3 lakh will be attained on 22 May 2020 and 26 May 2020, respectively.

## Introduction

COVID-19 has gripped the world and is spreading at such a speed with more potency that prevention, mitigation and control measures are falling flat. Although recoveries are reported, but the pandemic is still not under control. The current article is also an attempt to predict the spread of COVID-19 in India from the available statistical data and tools.

## Global Prediction Attempts

Many researchers tried to understand the pattern of spread, based on available data. In China one model is developed to map the spread of COVID-19 and the same is assumed to be accurate enough for both backward confirmation and forward extrapolation^i^. In yet another article, mathematical tools are exploited to understand the behaviour of infections and establishment of prediction tools in case of epidemics^ii^. The prediction strategy is also extended to spread of COVID-19 in Western World and major parameters like turning point, attack rate and durations are captured through modelling^iii^. The paper discusses 6 countries including France, USA, Canada, Italy, Germany and UK. It is predicted that a turning point of 69 days and recovery by first week of June 2020. Overall, data available is modelled by various exponential factors and the same is used to predict the nature of disease in future. However, a realistic and deterministic prediction strategy in Indian Context is missing and is not available. This paper is an attempt to predict the outburst of COVID-19 in India using exponential model without any logistic control support. Currently, the model is developed for rising trend and control is not assumed to be taking place in near future. The paper predicts infections of COVID-19 in India, for next one month using developed model.

## Current Trend in India

The confirmed cases for India, are taken from the website, earmarked for reporting confirmed cases of COVID-19 for all the regions, country and states of the world^iv^. The data is compiled from 15 March 2020 till 30 April 2020 for daily increase in number of confirmed cases of COVID-19 in India. The variation is plotted to understand the trend (Figure 1). As indicated in the figure, on 30 April 2020, 1823 new confirmed cases are added to the already existing confirmed cases of above 33000. Although there was a spike on 26 April 2020 (Sunday), but there is continuous rise there onwards.

**Figure 1.**
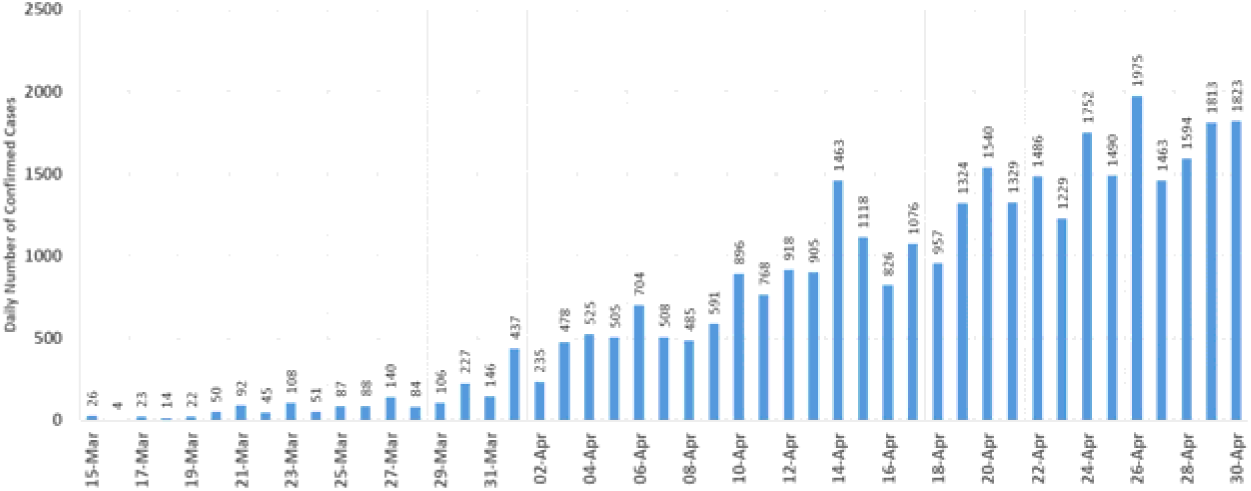
Daily Increase in Number of Confirmed Cases in India.

It is clear that the daily variation has been affected significantly by many factors including lockdown, bulk infection, public outbursts, government directives, people response, state-level implementation and so on. So, this cannot be fitted into any mathematical model for further analysis. The data is compiled for cumulative values of confirmed cases from the same site. As far as variation of cumulative confirmed cases in India from 15 March 2020, onwards is concerned, it has been on the rise consistently (Figure 2). It is looking like a smooth variation with monotonous rise. While considering cumulative figures, the occasional spikes are not clearly visible. So, this graph can be taken to predict the variation, for ease of mathematical modelling.

**Figure 2.**
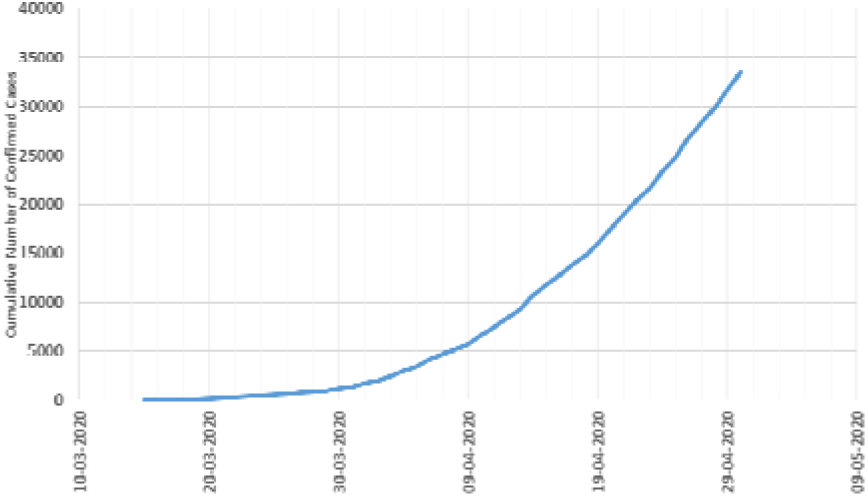
Cumulative Confirmed Cases of COVID-19 in India.

However, it is clear that the daily rise or variation is a better data for modelling and trend on the basis of data of previous dates may be helpful in prediction. Although, mathematically complicated, the daily variation of confirmed cases are taken for analysis, modelling and prediction. The data for two of the states of India, where confirmed cases are rising at a faster pace is considered. They are shown as Figure 3. A sudden spike in both the cases at around 18 April 2020 is obvious from the graph.

**Figure 3.**
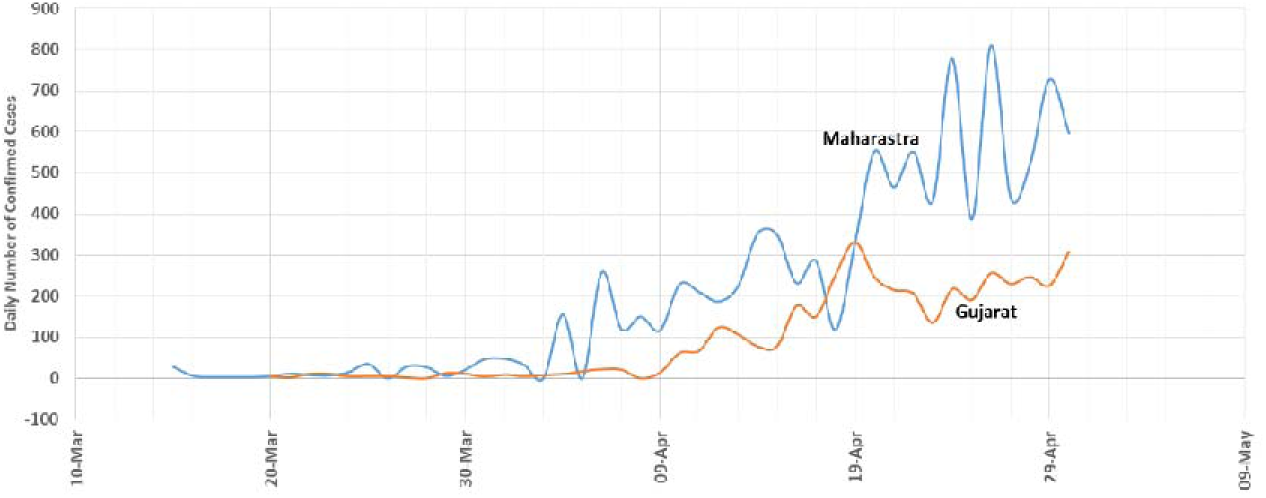
Daily COVID-19 Infections at Maharastra and Gujarat.

For completeness, some more states with high daily infection rates are taken, so that the mathematical model can be validated with them. The data of Delhi, Madhya Pradesh, Rajasthan and Uttar Pradesh is taken. They are assumed to be representing infection trends of all the states in India (Figure 4). It is observed that Delhi is showing many spikes and variation is not at all consistent. Delhi showed sudden burst of cases, as depicted by upward spikes on 4 April, 10 April, 12 April, 14 April, 19 April and 27 April. Compared to that Madhya Pradesh has relatively smooth variation, but has upward spikes on 11 April, 15 April, 17 April and 24 April. Rajasthan has further consistent data with upward spikes on 12 April and 22 April. Unfortunately, the dates of upward spikes in number of confirmed cases are not the same in any two states. Uttar Pradesh showed an upward spike on 14 April, which matches with one of the spikes of Delhi.

**Figure 4.**
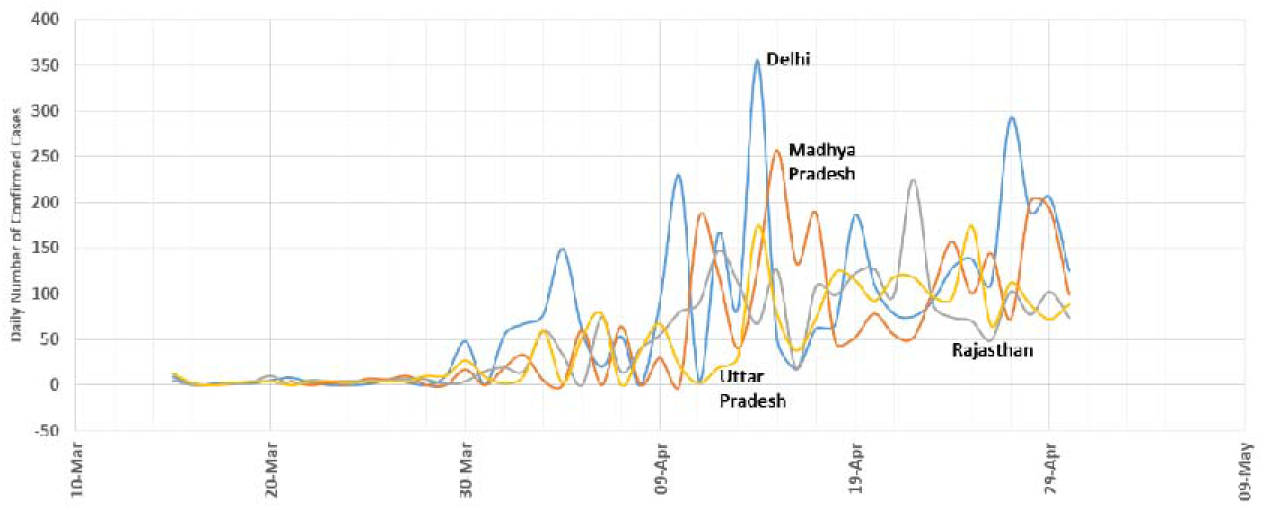
Daily variation of 4 More States in India.

## Prediction Strategy

For predicting spread of pandemics, mathematical tools are implemented earlier also. It is reported that Daniel Bernoulli, mathematician tried to map the spread of smallpox in 18^th^ century. Similarly statistical tools and mapping were used in next century for predicting Cholera spread. Attempts are also made to COVID-19 prediction. As per prediction on 24 March 2020, by Centre for Disease Dynamics, Economics and Policy predicted 3-4 Crore infections of mild COVID-19 in India by July 2020, which can be delayed by lockdown and only minor change in number was expected. Similarly, on 22 March 2020 COV-IND-19 study group predicted, 161 per lakh population by 15^th^ May 2020. This amounts to 22 lakh cases in total without any control measures. However, the preventive measures can reduce the same to 1 per lakh, making it a mere 13800. Currently both the numbers are off by large amount and some intermediate strategy is to be adopted for correct prediction based on current speed of spread, assuming the rate will continue in same manner and opening lockdown is not affecting the scenario to a sudden large outburst.

State wise data is compiled and studied with various mathematical models to understand the behaviour, assuming that the changes due to sudden outbursts are natural phenomena and not the isolated cases. It is also assumed that similar sudden outbursts may be possible in future also, due to attributable reasons. In developing a prediction strategy, following features are indirectly playing role:-(i) Infection spread rate, (ii) Recovery/Cure rate, (iii) Hidden outburst factor. But the confirmed cases are taken, which in combined way, takes care of all variations. The prediction strategy identifies a population growth model, which is applicable for the rise of microorganisms. It is assumed that there is no logistic model strategy, because currently the rise rate of infection is increasing and there is no slowing down, observed in the real data.

Since population is different in different areas and India shows a combination of all the states, it is assumed to rationalize the number of cases by taking Gujarat as reference. The ratio of population to current infection for each state and India is maintained same and daily infection for all states and India is rationalized to the population of Gujarat. The modified curve is shown as Figure 5.

**Figure 5.**
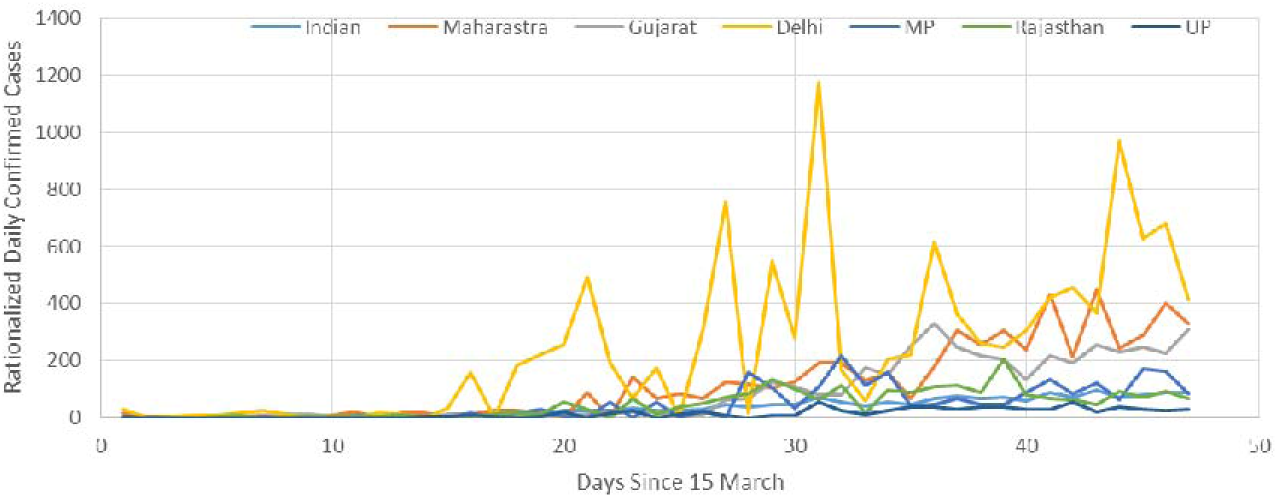
Rationalized Data for 6 States of India and India.

The data clearly indicates that except for the three states, Delhi, Maharastra and Gujarat, other states are following almost similar trend. The rationalized number of infections are lower in other states and accordingly, the predictions will have a slower rise in other states. An attempt is made to fit the same exponential model for all the cases, with proper constants. The Model used for prediction is given below

> H(t) = H(0) x Exp[s x t] + B, where t = time elapsed from reference (15 March 2020 in this case), H(0) = number of infections on 15 March 2020, H(t) = Rationalized infection at time t, s = intrinsic rate of increase, B = Offset value for daily infections (to cater for sudden outburst).

The major assumptions made are the constant are independent of space and time and are virtually invariant. All infectious microorganisms have equal potency to spread and it is independent of time lapsed and region. The rate of infection is continuous and not discrete, except for the sudden outburst due to specific reasons. There is no logical control model is applied, at present because plateau in rising trend has not been achieved yet.

## Prediction

It is clear that three states, Delhi, Maharastra and Gujarat has very high growth rate for the rationalized data. Same exponential model with different constants are fitted for the entire population and in the exponential model, the initial infections is changed to fit the proper curve. The modelled data for the 3 states and India is shown as Figure 6. Despite variation the curve can assumed to be a good fit.

**Figure 6.**
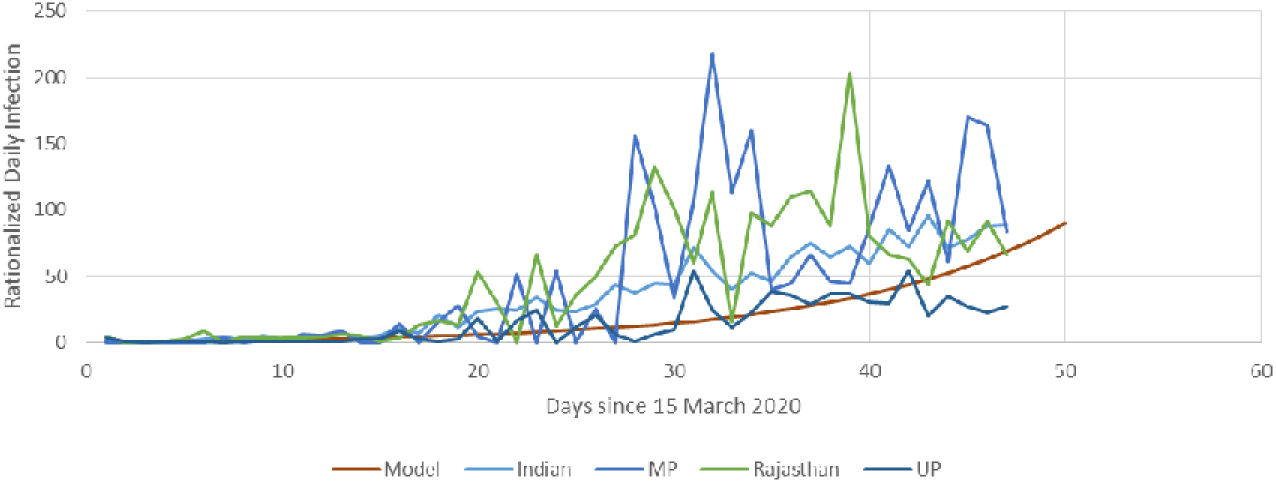
Modeling of Rationalized Daily Infection.

However, the same data is not applicable to other states and for them the initial value of infected case is taken higher and further data is fitted. The model predicts that in these three cases, Maharastra, Gujarat and Delhi, the initial infection were 2.5 times higher than others. It indirectly says that the rate of rise of infection is same in all the states, but in these three states the value at initial time was higher, which resulted in higher growth, currently. The modelling for these three states is given as Figure 7. Slight upward offset is also applied, to cater for the sudden local time-dependent outbursts, in these areas, which will nullify as time progresses. It is also observed that such outbursts are not reflected in the overall daily infection of India, because of larger population domain. Based on the developed exponential model, prediction is made for next one month at an interval of 5 days and the values are tabulated below for all the states and India.

**Figure 7.**
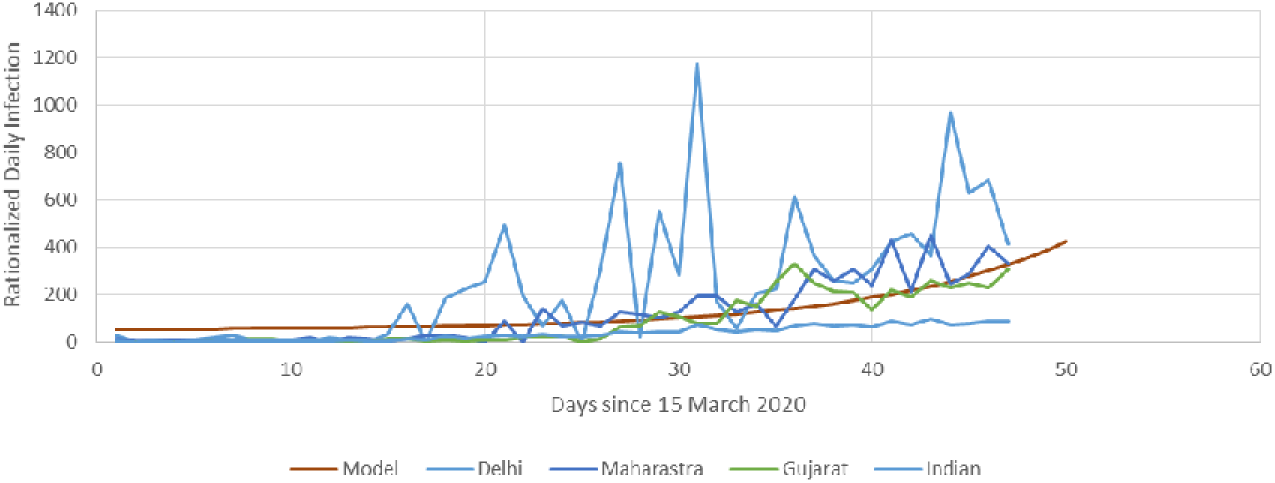
Modeling for High Infection States.

**Table 1.** Prediction of Daily Infection till 30 May 2020.

| Date | India | Maharastra | Gujarat | Delhi | MP | Rajasthan | UP |
| --- | --- | --- | --- | --- | --- | --- | --- |
| 30-Apr-20 | 2258 | 590 | 325 | 98 | 129 | 122 | 351 |
| 05-May-20 | 3722 | 913 | 503 | 152 | 213 | 201 | 578 |
| 10-May-20 | 6137 | 1447 | 797 | 241 | 352 | 332 | 953 |
| 15-May-20 | 10118 | 2326 | 1282 | 388 | 580 | 547 | 1571 |
| 20-May-20 | 16681 | 3776 | 2081 | 630 | 956 | 901 | 2591 |
| 25-May-20 | 27502 | 6168 | 3399 | 1028 | 1576 | 1486 | 4272 |
| 30-May-20 | 45344 | 10110 | 5571 | 1685 | 2598 | 2449 | 7043 |

Based on this calculation, the prediction for India is plotted as Figure 8. The variation is found to be matching to the current confirmed cases, as on 30 April 2020. In absence of correct controlling factors, the cases are likely to cross 1 lakh mark on 15 May 2020. It will have further rise at a very fast pace, due to exponential effect of the model. The prediction calculated 2 lakh mark on 22 May 2020 and 3 lakh by 26 May 2020. However, it is expected that some decaying factor will become prevalent including immunity, environment, awareness and mitigation effects, which prevents the exponential rise as per model.

**Figure 8.**
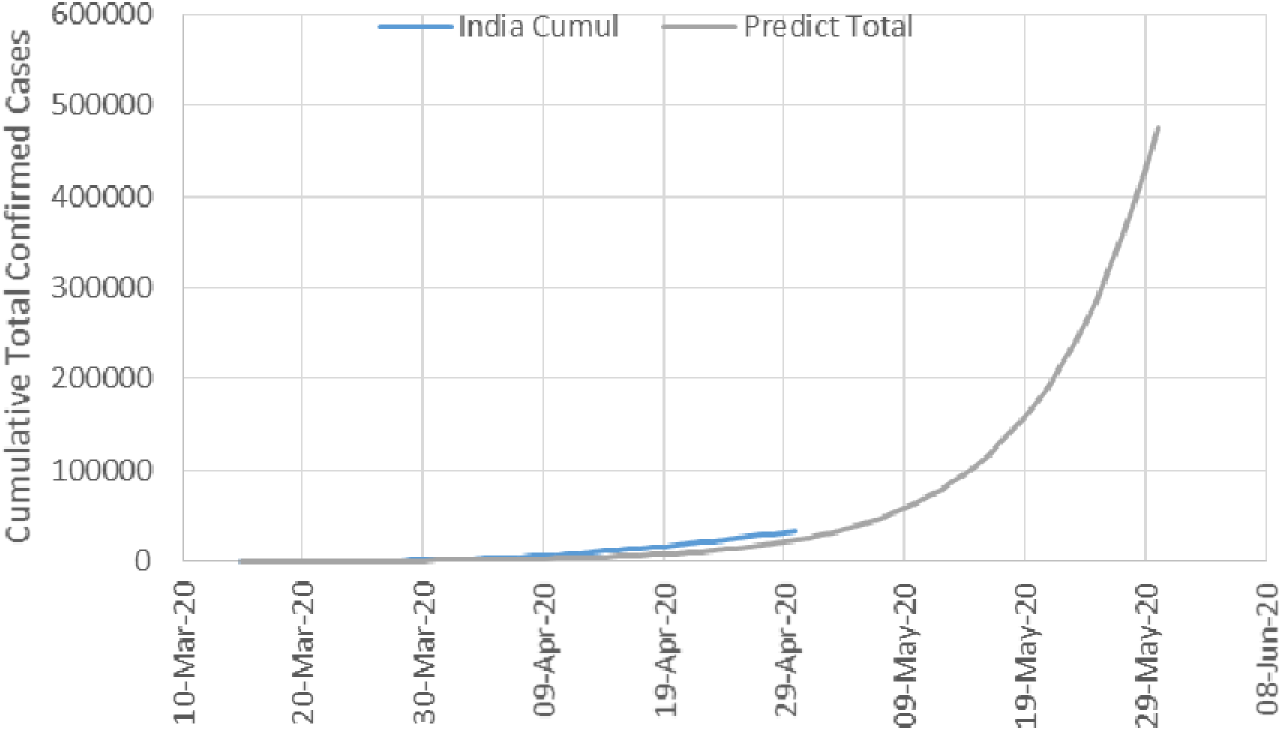
Prediction for Indio against Available Data.

## Conclusion

The daily infection of COVID-19 in India and 6 of its states are modelled, based on exponential growth. India on an average has a slower rate of rise, as compared to three states namely Maharastra, Gujarat and Delhi. However, most of the states of India behaves in same way as India. These three states also exhibited same rate of growth, but the initial number of infections can be assumed to be 2.5 times others, due to sudden outbursts. The logistic factor is not applied, as the rate of rise of daily infections have rising trend at present. If the case proceeds in same manner, then number of infections in India will reach 1 lakh mark by 15 May 2020.

Author Dr Himanshu Shekhar is working as Director(Admin) in the office of Director General, DRDO at Pune, India and has been associated with development of Indigenous products for Defence. He has 26 books and more than 120 technical research publications in international and national journals and conferences. He is the recipient of Young Scientist Award, Mr Engineer Award, Agni Award for Excellence in Self Reliance and national science day oration award. He has undergone 3 basic training courses on COVID-19 through Integrated Government Online Training (IGOT) and has been monitoring the spread of COVID-19, closely.

## Data Availability

The data is collected from Googlenews about status of daily infections in India and the same is mentioned as one of the references

## Reference

i Lixiang Li, Zihang Yang, Zhongkai Dang, Cui Meng, Jingze Huang, Haotian Meng, Deyu Wang, Guanhua Chen Jiaxuan Zhang, Haipeng Peng, Yiming Shao, Propagation Analysis and Prediction of the COVID-19, Infectious Disease Modelling, Volume 5, 2020, Pages 282–292

ii Julien Arino, Mathematical Epidemiology in Data Rich World, Infectious Disease Modelling, Volume 5, 2020, Pages 161–168.

iii Xiaolei Zhang, Renjun Ma, Lin Wang, Predicting turning point, duration and attack rate of COVID-19 outbreaks in major Western countries, Chaos, Solition and Fractals, 135, 2020, 109829.

iv https://news.google.com/covid19/map?hl=en-IN&gl=IN&ceid=IN:en

